# Natural History and Post-transplant Outcomes Among Patients with Kidney Leukocyte Cell-Derived

**DOI:** 10.64898/2026.07.31.26359278

**Authors:** Iqra Fatima Munawar Ali, Pedro A. Lopez-Gutierrez, Nimra Iftikhar, Valeria Hanson, Shumaila Afrin, Lorena Saelices Gomez, Pablo Garcia, Christos Argyropoulos

## Abstract

**Introduction:** ALECT2 is the third most common cause of renal amyloidosis in the United States, with a strong predilection for Hispanic patients in the Southwest. (PMID:24522497) Despite its regional prevalence, data on the trajectory of kidney function decline and post-transplant outcomes remain poorly characterized.

**Methods:** This is a retrospective study of patients diagnosed with ALECT2 by kidney biopsy at UNM from January 2014 to April 2024. Pre- and post-biopsy eGFR trajectories and post-transplant allograft function were analyzed.

**Results:** Twelve patients had pre-biopsy data available, with a mean age at diagnosis of 64 ± 12.0 years; 8 (67%) were female, and 8 identified as Hispanic. Mean eGFR at diagnosis was 31 (SD 29), indicating advanced kidney disease, with 67% diagnosed as CKD stage 4 or 5. Concomitant kidney pathology was identified in 50% of patients, with diabetic nephropathy present in two-thirds of those cases. A significant change in the eGFR trajectory was identified at the time of biopsy through change-point analysis [Figures A, C, D]. Among the four patients who underwent kidney transplantation, allograft eGFR remained stable over more than 60 months of follow-up [Figure D].

**Conclusion:** In this cohort of predominantly Hispanic patients, ALECT2 amyloidosis presented with advanced CKD and accelerated loss of kidney function, which prompted a biopsy that detected the disease. Post-transplant allograft function was well maintained, supporting kidney transplantation as a viable treatment strategy for ALECT2-associated kidney disease.

## INTRODUCTION

Leukocyte cell-derived chemotaxin 2 (LECT2) has emerged as the third most common cause of renal amyloidosis in the United States, accounting for 2.5–2.7% of cases in large biopsy series.(1,2) In the Southwest United States ALECT2 accounts for up to 54% of all renal amyloid cases, and autopsy studies have identified the disease in 3.1% of Hispanic adults in New Mexico compared to 0.7% of non-Hispanic Caucasians.(3) Beyond the Hispanic population ALECT2 has been increasingly recognized across various other ethnic groups.(4)

Despite its high prevalence, the current literature reports outcomes exclusively from the time of biopsy onwards, leaving the degree and duration of diagnostic delay unquantified. No disease-specific treatment has been identified, transplant outcome data remain sparse, and no dedicated outcomes series exists from the Southwest United States, one of the highest prevalence regions globally. Herein we present a biopsy-diagnosed LECT 2 cohort with longitudinal kidney function data extending up to 60 months prior to diagnosis, post-transplant allograft outcomes, and change point analysis to identify the timing and magnitude of change in kidney function trajectory around the time of diagnostic biopsy.

## METHODS

### Study Population

We performed a retrospective analysis of patients diagnosed with LECT2 amyloidosis at the University of New Mexico Hospitals via native kidney biopsy with positive LECT2 staining.

Baseline characteristics were taken within a window of ±90 days relative to the date of the diagnostic kidney biopsy and were extracted from the electronic medical record. (Table 1)

Estimated Glomerular Filtration Rate (eGFR) was calculated for all time points using the CKD-EPI 2021 race free creatinine equation.

## RESULTS

A total of 12 subjects were identified, with the mean age at diagnosis 64± 12.0 years, 8(67%) were females, and 8 identified as Hispanic. Mean eGFR at diagnosis was 31 (SD 29) indicating advanced kidney disease with 67% diagnosed as CKD stage 4 or 5. Concomitant kidney pathology was identified in 50% with diabetic nephropathy being present in about 66% of those. A significant change in the eGFR trajectory identified at the time of biopsy through change point analysis [Figures A,C,D] Among four patients who underwent kidney transplantation, allograft eGFR remained stable over more than 60 months of follow-up [Figure D], allograft eGFR remained stable over more than 60 months of follow-up without evidence of disease recurrence. Uniquely, a significant change in the eGFR trajectory is identified at the time of biopsy through change point analysis, with significant change in the before and after biopsy data.

## DISCUSSION

LECT2 is a multifunctional protein produced mainly by hepatocytes, and implicated in diverse biological processes including liver regeneration, immune modulation, bone metabolism, glucose homeostasis, and oncogenesis. (2,5) ALECT2 typically presents in older adults with progressive renal impairment and variable degrees of proteinuria. (3) Clinically, ALECT-2 presents without cardiac involvement unlike other amyloidosis subtypes. (6)

While Said et al. established serum creatinine of 2.0 mg/dL as the critical threshold beyond which progression to ESRD becomes significantly more likely, (6) our change point analysis demonstrates a significant shift in eGFR trajectory around the time of diagnostic biopsy, suggesting that substantial kidney function loss has already occurred before ALECT2 is identified.

Among four patients who underwent kidney transplantation, in our set-up, allograft eGFR remained stable over more than 60 months of follow-up without evidence of disease recurrence, extending beyond the 20-month follow-up reported by Said et al., where disease recurred in one of five recipients without graft loss. (6) Hence, our study is offering a distinct chance to unravel the natural history of ALECT2 in the transplanted kidney. The extended follow-up in our transplanted patients may provide insight into both the pathogenesis of ALECT2 and its natural history after transplantation, including whether allograft involvement develops over time and whether recurrence is associated with progressive graft dysfunction. Together these findings suggest that kidney transplantation is a viable treatment option for eligible patients with ALECT2-associated renal failure.

This study has several limitations. Our cohort is small and from a single center, which may limit generalizability; however given that the Southwest United States has one of the highest prevalence of ALECT2 globally, our findings remain relevant. The change point represents a visual assessment rather than a formal statistical analysis, and confounding by indication is unavoidable as patients with more rapidly declining kidney function are more likely to undergo biopsy sooner. This highlights the need for better diagnostic tools to identify patients earlier before irreversible damage occurs. In terms of transplant outcomes, our findings along with those of Said et al. support kidney transplantation as a viable option for ALECT2-associated end stage renal disease. However given the small number of transplanted patients across all published studies, longer follow up and larger studies are needed to better understand post-transplant outcomes and disease recurrence.

**Figure 1.**
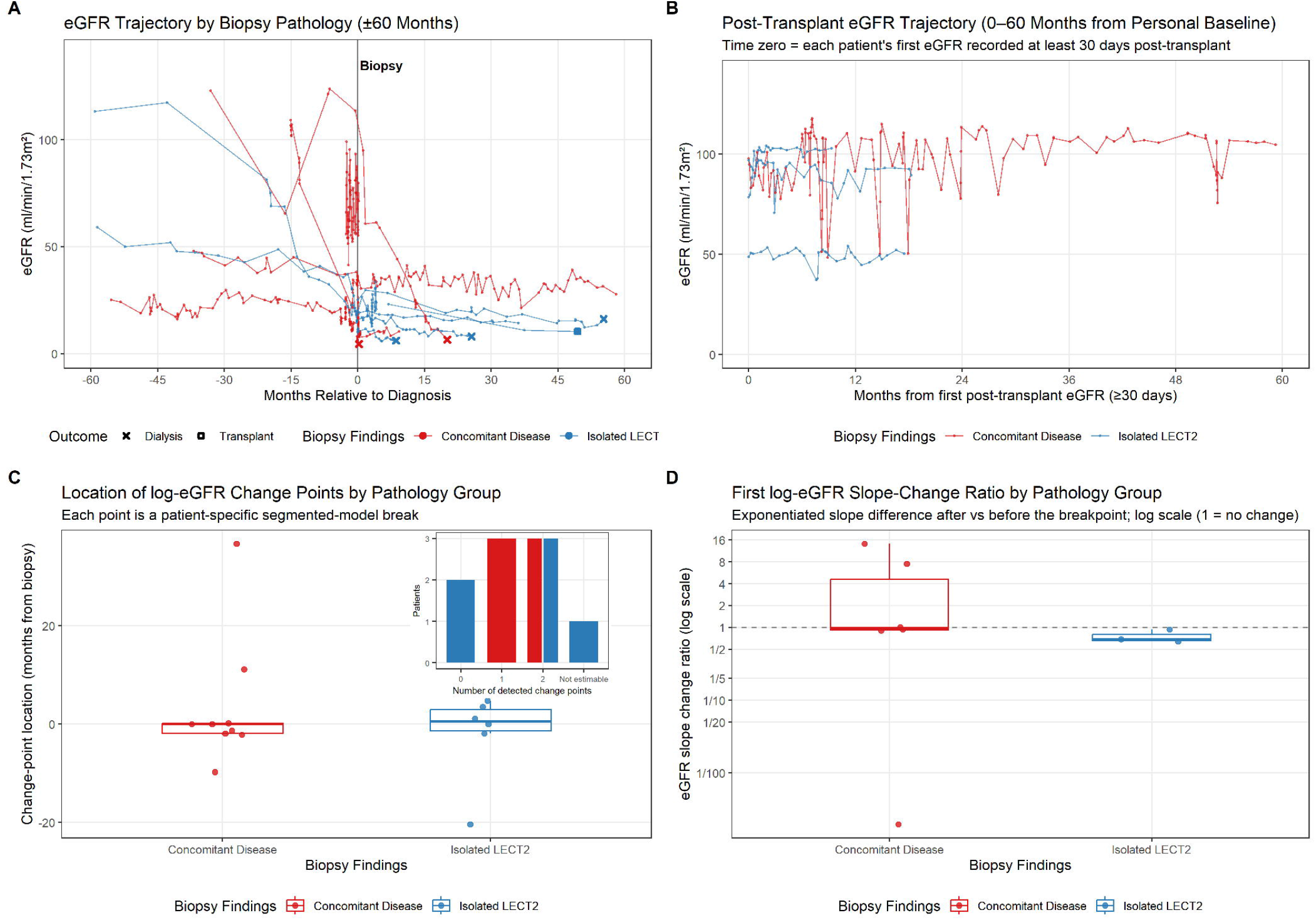

## Supporting information

https://docs.google.com/document/d/1t1cK6JqyhtJGXGZrHK0fLV-BupRB4RWe/edit?usp=sharing&ouid=115946396145860040621&rtpof=true&sd=true

## Data Availability

All data produced in the present study are available upon reasonable request to the corresponding author.

## Disclosure

L.S. is a co-founder of AmyGo and reports consulting and/or advisory board fees from Pfizer, AstraZeneca, and AmyGo, and research support from the NIH, AstraZeneca, and UT Southwestern Medical Center.

## Notes

### Competing Interest Statement

The authors have declared no competing interest.

### Author Declarations

The Human Research Protection Office of the Health Sciences Center at the University of New Mexico gave ethical approval for this work. Protocol number 19-351. Informed consent was waived as this was a retrospective study.

