## Supplementary material for "Natural History and Post-transplant Outcomes Among Patients with Kidney Leukocyte Cell-Derived": https://docs.google.com/document/d/1t1cK6JqyhtJGXGZrHK0fLV-BupRB4RWe/edit?usp=sharing&ouid=115946396145860040621&rtpof=true&sd=true

Table 1: Baseline Characterstics

| Characteristic | N = 12[1] |
|---|---|
| **Age at biopsy (years)** | |
| Mean (SD) | 64 (12) |
| Median (Q1, Q3) | 66 (55, 74) |
| **Gender** | |
| Female | 8 (67%) |
| Male | 4 (33%) |
| **Race** | |
| Native American | 2 (29%) |
| White | 5 (71%) |
| (Missing) | 5 |
| **Ethnicity** | |
| Hispanic | 8 (100%) |
| (Missing) | 4 |
| **HTN - Pre-biopsy** | |
| 0 | 6 (50%) |
| 1 | 6 (50%) |
| **HTN - Post-biopsy** | |
| 0 | 5 (42%) |
| 1 | 7 (58%) |
| **DM - Pre-biopsy** | |
| 0 | 6 (50%) |

| Characteristic | N = 12[1] |
|---|---|
| 1 | 6 (50%) |
| **DM - Post-biopsy** | |
| 0 | 6 (50%) |
| 1 | 6 (50%) |
| **Liver Disease - Pre-biopsy** | |
| 0 | 10 (83%) |
| 1 | 2 (17%) |
| **Liver Disease - Post-biopsy** | |
| 0 | 10 (83%) |
| 1 | 2 (17%) |
| **ACEi/ARB - Pre-biopsy** | |
| 0 | 8 (67%) |
| 1 | 4 (33%) |
| **ACEi/ARB - Post-biopsy** | |
| 0 | 8 (67%) |
| 1 | 4 (33%) |
| **SGLT2i - Pre-biopsy** | |
| 0 | 11 (92%) |
| 1 | 1 (8.3%) |
| **SGLT2i - Post-biopsy** | |
| 0 | 11 (92%) |

| Characteristic | N = 12[1] |
|---|---|
| 1 | 1 (8.3%) |
| **Concomitant Kidney Pathology** | |
| IgA Nephropathy | 1 (8.3%) |
| Membranous nephropathy with IgG kappa deposits | 1 (8.3%) |
| Nodular Diabetic Glomerulosclerosis | 4 (33%) |
| None | 6 (50%) |
| **CKD Stage** | |
| G1 (>90) | 1 (8.3%) |
| G2 (60-89) | 0 (0%) |
| G3a (45-59) | 1 (8.3%) |
| G3b (30-44) | 2 (17%) |
| G4 (15-29) | 5 (42%) |
| G5 (<15) | 3 (25%) |
| **Proteinuria category (UPCR)** | |
| < 0.5 g/g | 3 (25%) |
| 0.5 - 1.0 g/g | 3 (25%) |
| > 1.0 g/g | 6 (50%) |
| **Ended on dialysis** | 5 (42%) |
| **Days from biopsy to dialysis** | |
| Mean (SD) | 728 (735) |
| Median (Q1, Q3) | 628 (282, 794) |

| Characteristic | N = 12[1] |
| --- | --- |
| (Missing) | 7 |
| **Had transplant** | 1 (8.3%) |
| **Days from biopsy to transplant** | |
| Mean (SD) | 2,028.0000 (NA) |
| Median (Q1, Q3) | 2,028.0000 (2,028.0000, 2,028.0000) |
| (Missing) | 11 |
| **Creatinine** | |
| Mean (SD) | 3.05 (2.21) |
| Median (Q1, Q3) | 2.66 (1.88, 3.40) |
| **Days to Creatinine from biopsy** | |
| Mean (SD) | 16 (62) |
| Median (Q1, Q3) | 0 (-4, 2) |
| **eGFR** | |
| Mean (SD) | 31 (29) |
| Median (Q1, Q3) | 21 (15, 36) |
| **Days to eGFR from biopsy** | |
| Mean (SD) | 16 (62) |
| Median (Q1, Q3) | 0 (-4, 2) |
| **UPCR** | |
| Mean (SD) | 4.1 (6.0) |
| Median (Q1, Q3) | 1.0 (0.4, 4.3) |
| **Days to UPCR from biopsy** | |

| Characteristic | N = 12[1] |
|---|---|
| Mean (SD) | 42 (79) |
| Median (Q1, Q3) | 4 (-6, 46) |
| **Albumin** | |
| Mean (SD) | 3.05 (0.84) |
| Median (Q1, Q3) | 3.20 (2.45, 3.75) |
| **Days to Albumin from biopsy** | |
| Mean (SD) | 29 (72) |
| Median (Q1, Q3) | 7 (-1, 47) |
| **ACR** | |
| Mean (SD) | 2,563 (4,711) |
| Median (Q1, Q3) | 773 (248, 2,248) |
| (Missing) | 4 |
| **Days to ACR from biopsy** | |
| Mean (SD) | 148 (201) |
| Median (Q1, Q3) | 151 (2, 207) |
| (Missing) | 4 |
| **AlkPhos** | |
| Mean (SD) | 107 (45) |
| Median (Q1, Q3) | 95 (67, 144) |
| **Days to AlkPhos from biopsy** | |
| Mean (SD) | 30 (73) |
| Median (Q1, Q3) | 10 (-3, 59) |

| Characteristic | N = 12[1] |
|---|---|
| **ALT** | |
| Mean (SD) | 24 (15) |
| Median (Q1, Q3) | 24 (17, 25) |
| **Days to ALT from biopsy** | |
| Mean (SD) | 30 (73) |
| Median (Q1, Q3) | 10 (-3, 59) |
| **AST** | |
| Mean (SD) | 21 (10) |
| Median (Q1, Q3) | 18 (14, 26) |
| **Days to AST from biopsy** | |
| Mean (SD) | 30 (73) |
| Median (Q1, Q3) | 10 (-3, 59) |
| **Bicarbonate** | |
| Mean (SD) | 21.0 (4.1) |
| Median (Q1, Q3) | 20.5 (18.5, 25.0) |
| **Days to Bicarbonate from biopsy** | |
| Mean (SD) | 16 (62) |
| Median (Q1, Q3) | 0 (-4, 2) |
| **BUN** | |
| Mean (SD) | 47 (22) |
| Median (Q1, Q3) | 47 (32, 54) |
| **Days to BUN from biopsy** | |

| Characteristic | N = 12[1] |
|---|---|
| Mean (SD) | 16 (62) |
| Median (Q1, Q3) | 0 (-4, 2) |
| **Calcium** | |
| Mean (SD) | 8.63 (0.61) |
| Median (Q1, Q3) | 8.50 (8.20, 9.00) |
| **Days to Calcium from biopsy** | |
| Mean (SD) | 16 (62) |
| Median (Q1, Q3) | 0 (-4, 2) |
| **Chloride** | |
| Mean (SD) | 108.33 (3.31) |
| Median (Q1, Q3) | 109.50 (105.00, 111.50) |
| **Days to Chloride from biopsy** | |
| Mean (SD) | 16 (62) |
| Median (Q1, Q3) | 0 (-4, 2) |
| **Cholesterol** | |
| Mean (SD) | 177 (47) |
| Median (Q1, Q3) | 181 (141, 201) |
| (Missing) | 2 |
| **Days to Cholesterol from biopsy** | |
| Mean (SD) | 458 (1,062) |
| Median (Q1, Q3) | 63 (-57, 395) |
| (Missing) | 2 |

| Characteristic | N = 12[1] |
|---|---|
| **DirectBili** | |
| Mean (SD) | 0.19 (0.17) |
| Median (Q1, Q3) | 0.15 (0.10, 0.20) |
| **Days to DirectBili from biopsy** | |
| Mean (SD) | 29 (73) |
| Median (Q1, Q3) | 10 (-6, 59) |
| **Globin** | |
| Mean (SD) | 3.9000 (NA) |
| Median (Q1, Q3) | 3.9000 (3.9000, 3.9000) |
| (Missing) | 11 |
| **Days to Globin from biopsy** | |
| Mean (SD) | 104.0000 (NA) |
| Median (Q1, Q3) | 104.0000 (104.0000, 104.0000) |
| (Missing) | 11 |
| **HDL** | |
| Mean (SD) | 55 (19) |
| Median (Q1, Q3) | 46 (43, 79) |
| (Missing) | 2 |
| **Days to HDL from biopsy** | |
| Mean (SD) | 458 (1,062) |
| Median (Q1, Q3) | 63 (-57, 395) |
| (Missing) | 2 |

| Characteristic | N = 12[1] |
| --- | --- |
| **Hemoglobin** | |
| Mean (SD) | 11.33 (2.28) |
| Median (Q1, Q3) | 10.95 (9.25, 13.45) |
| **Days to Hemoglobin from biopsy** | |
| Mean (SD) | 18 (61) |
| Median (Q1, Q3) | 0 (0, 2) |
| **IndirectBili** | |
| Mean (SD) | 0.45 (0.48) |
| Median (Q1, Q3) | 0.30 (0.20, 0.50) |
| **Days to IndirectBili from biopsy** | |
| Mean (SD) | 340 (882) |
| Median (Q1, Q3) | 35 (-6, 247) |
| **LDL** | |
| Mean (SD) | 88 (44) |
| Median (Q1, Q3) | 83 (49, 123) |
| (Missing) | 2 |
| **Days to LDL from biopsy** | |
| Mean (SD) | 458 (1,062) |
| Median (Q1, Q3) | 63 (-57, 395) |
| (Missing) | 2 |
| **Microalbumin** | |
| Mean (SD) | 79 (155) |

| Characteristic | N = 12[1] |
|---|---|
| Median (Q1, Q3) | 1 (1, 157) |
| (Missing) | 8 |
| **Days to Microalbumin from biopsy** | |
| Mean (SD) | 2,333 (1,731) |
| Median (Q1, Q3) | 2,681 (1,068, 3,598) |
| (Missing) | 8 |
| **Plts** | |
| Mean (SD) | 266 (101) |
| Median (Q1, Q3) | 262 (198, 315) |
| **Days to Plts from biopsy** | |
| Mean (SD) | 17 (61) |
| Median (Q1, Q3) | 1 (-1, 5) |
| **Potassium** | |
| Mean (SD) | 4.32 (0.74) |
| Median (Q1, Q3) | 4.40 (3.55, 4.90) |
| **Days to Potassium from biopsy** | |
| Mean (SD) | 16 (62) |
| Median (Q1, Q3) | 0 (-4, 2) |
| **Protein** | |
| Mean (SD) | 6.75 (1.16) |
| Median (Q1, Q3) | 6.90 (5.45, 7.70) |
| **Days to Protein from biopsy** | |

| Characteristic | N = 12[1] |
|---|---|
| Mean (SD) | 30 (73) |
| Median (Q1, Q3) | 10 (-3, 59) |
| **Sodium** | |
| Mean (SD) | 138.58 (3.63) |
| Median (Q1, Q3) | 139.00 (136.00, 141.00) |
| **Days to Sodium from biopsy** | |
| Mean (SD) | 16 (62) |
| Median (Q1, Q3) | 0 (-4, 2) |
| **TotalBili** | |
| Mean (SD) | 0.63 (0.64) |
| Median (Q1, Q3) | 0.50 (0.30, 0.65) |
| **Days to TotalBili from biopsy** | |
| Mean (SD) | 30 (73) |
| Median (Q1, Q3) | 10 (-3, 59) |
| **Triglyceride** | |
| Mean (SD) | 165 (84) |
| Median (Q1, Q3) | 140 (110, 231) |
| (Missing) | 2 |
| **Days to Triglyceride from biopsy** | |
| Mean (SD) | 458 (1,062) |
| Median (Q1, Q3) | 63 (-57, 395) |

| Characteristic | N = 12[1] |
|---|---|
| (Missing) | 2 |
| **WBC** | |
| Mean (SD) | 8.42 (2.38) |
| Median (Q1, Q3) | 9.10 (6.20, 10.05) |
| **Days to WBC from biopsy** | |
| Mean (SD) | 18 (61) |
| Median (Q1, Q3) | 1 (-1, 7) |

[1]n (%)
